# Using novel internet-based sources for Population Size Estimation of Gay, bisexual, and other cisgender men who have sex with men

**DOI:** 10.64898/2026.09.14.26363087

**Authors:** Yuanqi Mi, Alex Garner, Maguette Niang, Chenglin Hong, Sean Howell, Jody Herman, Ian Holloway, Stefan Baral

## Abstract

Population size estimates (PSE) for gay, bisexual, and other cisgender men who have sex with men (GBMSM) remain difficult where stigma and criminalization limit data collection and HIV resource allocation. We aim to assess whether Google Trends and a gay social networking app can provide standardized, low-cost population size estimates (PSE) for gay, bisexual, and other cisgender men who have sex with men (GBMSM) in Senegal, Ghana, Mozambique, and Indonesia.

We analyzed country-level and local GT relative search volumes for “porn” and “gay porn” from September 2024 to September 2025. Pornhub global review data were applied to produce proportional GBMSM PSEs. App estimates were derived from monthly active users and extrapolated to annual users. Proportional estimates were multiplied by the male population aged 15–49 years to obtain absolute PSE and compared with UNAIDS Key Population Atlas estimates. Sensitivity analyses used survey-based multipliers and alternative search-term combinations.

GTPSE ranged from 0.37% in Mozambique to 1.92% in Indonesia, while app-based estimates ranged from 0.29% in Mozambique to 1.34% in Ghana. GTPSE exceeded UNAIDS estimates in Ghana, Mozambique, and Indonesia. App-based estimates exceeded UNAIDS estimates in Ghana and Mozambique. Maputo’s estimated GBMSM proportion was substantially higher than Mozambique’s national estimate.

GTPSE generally exceeded UNAIDS estimates, which may reflect underestimation in stigmatizing settings where GBMSM are less likely to be reached through field-based methods or mainstream health services. Social network app estimates were lower in some countries, highlighting variability in platform access and geographic differences in the concentration of social networks.

Internet-derived data may offer a low-cost, feasible approach to GBMSM PSE in low- and middle-income settings. Further validation through digital data linkages could strengthen HIV prevention and treatment planning.

**Author summary:** Reliable estimates of the number of gay, bisexual, and other cisgender men who have sex with men are important for HIV prevention and treatment services planning, but these estimates are often difficult to obtain in settings where stigma, criminalization, and limited resources constrain traditional field-based methods. In this study, we examined whether routinely available digital data could provide an alternative approach. We used Google Trends search data and aggregated data from a gay social networking app to estimate population sizes in Senegal, Ghana, Mozambique, and Indonesia, and compared with available UNAIDS estimates. We found that digital data sources produced broadly plausible but sometimes divergent estimates across countries and platforms. Google Trends estimates were generally higher than UNAIDS estimates. Social network app estimates were lower in some countries, highlighting variability in platform access and geographic differences in the concentration of social networks. Our findings suggest that digital data may complement existing population estimation methods, particularly where conventional approaches are difficult or costly to implement. Further validation across settings and data sources is needed before these methods can be used routinely for public health surveillance and service planning.

## Introduction

Gay, bisexual, and other cisgender men who have sex with men (GBMSM) have been shown to be disproportionately affected by HIV in every setting studied given intersecting individual, network-, and structural determinants(1). However, reliable data on GBMSM are often most limited in settings where stigma and criminalization are most pervasive and have historically constrained investment in HIV services. This is because individuals may be less likely to disclose same-sex practices or be reached through conventional in-person epidemiological studies or mainstream healthcare services. To inform evidence-based HIV responses, population size estimates (PSE) and the burden of HIV are needed to calculate the population attributable fraction to guide service delivery(2).

Empirically derived PSE typically rely on resource-intensive in-person methods including multisource capture–recapture, respondent-driven sampling successive sampling, among others(3). With increasing availability of digital data, novel data sources such as Google Trends (GT) and social networking apps represent potentially underutilized data to inform low-cost methodologies for GBMSM PSE(4). Characterizing valid strategies leveraging online platforms may represent sustainable strategies to inform estimations of HIV epidemic dynamics particularly in rights-constrained environments.

Existing UNAIDS PSE estimates vary substantially in methodological quality across countries, ranging from rigorous field-based studies to nationally submitted figures of uncertain provenance(5). In criminalized settings, national governments face structural political incentives to undercount GBMSM populations, raising questions about the reliability of official estimates as benchmarks in precisely the settings where GBMSM bear the greatest HIV burden. Previous digital approaches to GBMSM PSE leveraged Facebook interest targeting and gay social networking apps to generate estimates consistently higher than UNAIDS figures across 13 countries(4). However, Facebook discontinued detailed targeting of sensitive audience categories including sexual orientation in 2021-2022, eliminating this method. Simultaneously, the 2025 interruption of PEPFAR-funded programs has disrupted field-based PSE in many settings. These converging developments create an urgent need for standardized, low-cost digital PSE methods that are reproducible across countries, independent of national political pressures, and do not rely on platform data-sharing agreements that may be discontinued.

This study aimed to examine the utility of applying Google Trends (GT) internet search data to generate PSE (Google Trends Population Size Estimate [GTPSE]) for GBMSM and provide estimates of members of Grindr (a social app geared towards GBMSM) in four countries in Sub-Saharan Africa and Asia.

## Methods

We analyzed GT data and social networking app data from four countries supported by the US President’s Emergency Plan for AIDS Relief (PEPFAR): Senegal, Ghana, Mozambique, and Indonesia. These countries were selected because GBMSM PSE were needed for HIV programming and because each represents a setting where same-sex conduct is criminalized or heavily stigmatized, making official UNAIDS estimates particularly susceptible to political underreporting and field-based methods especially difficult to implement. Averaged weekly country-level and local GT relative search volumes for key terms “porn” and “gay porn” as a comparator term were examined from September 2024 to September 2025(6). According to Pornhub’s global review data, 64% of “porn” represents “men”(denominator) while 53%”gay porn” represents a subset of GBMSM (numerator)(7, 8). We used this approach to produce proportional GBMSM PSEs for country-level national estimates and local estimates (political or commercial capital city). The number of unique gay app monthly active users in 2021 who resided in each country were provided by a Gay Social Network app (Grindr) and were extrapolated to the number of annual users. Given the need to reliably estimate the proportion of GBMSM, these estimates were compared with most recent estimates from The Joint United Nations Programme on HIV/AIDS (UNAIDS) Key Population Atlas (where available)(3, 5). We applied internet access multipliers for Mozambique from its national household survey given limited access in the country. The primary app-based estimate was treated as an unadjusted app-reachable estimate among GBMSM. We multiplied the proportional GTPSE and social network app PSE values with the countries’ male adult population (15-49 years) according to the U.S. Census Bureau to obtain absolute PSE. We conducted internet surveys to generate survey-based multipliers for GBMSM who access porn and who use social networking apps among all GBMSM, which were applied as sensitivity analysis. Additional sensitivity analyses repeated the GTPSE calculations using “gay sex” relative to “sex” and “anal sex” relative to “sex” as alternative search-term combinations, applying the same gender and internet-access adjustments used in the primary analysis.

## Results

Across all four countries, GTPSE of MSM ranged from 0.37% (Mozambique) to 1.92% (Indonesia) of the adult male population, and social network app estimates ranged from 0.29% (Mozambique) to 1.34% (Ghana). Credibility intervals could not be computed due to the absence of absolute search volume data from Google Trends.

In three of four countries, GTPSE exceeded extrapolated UNAIDS estimates, with the largest discrepancies observed in Indonesia (GTPSE 1.92% vs UNAIDS 1.17%) and Ghana (GTPSE 1.18% vs UNAIDS 0.85%). Senegal was the notable exception, where GTPSE and UNAIDS estimates were approximately aligned (1.01% vs approximately 1.0%). Most subnational estimates in our sample were lower than or consistent with national estimates in three countries. In Mozambique, however, the estimated proportion in Maputo was substantially higher than the national estimate (2.49% vs. 0.37%). This is consistent with the greater concentration of GBMSM commonly observed in urban centers. (Table 1).

**Table 1.** Comparison of GBMSM estimates from UNAIDS KP Atlas, Social network app, and Google Trends.

| Country (local area*) | Relative national GTPSE, %, 2025 | Relative local GTPSE, %, 2025 | Absolute percentage difference national and local GTPSE, % , 2025 | National GTPSE***, 2025 | Social network app PSE****, 2025 | Social network app PSE, %, 2025 | UNAIDS KP Atlas, 2020 | UNAIDS estimates extrapolated for 2025 |
| --- | --- | --- | --- | --- | --- | --- | --- | --- |
| Senegal (Dakar) | 1.01 | 0.85 | 0.16 | 46,921 | 26,904 | 0.58 | 52,500 | 58,100 |
| Ghana (Accra) | 1.18 | 0.85 | 0.34 | 100,903 | 114,204 | 1.34 | 54,800 | 59,800 |
| Mozambique (Maputo**) | 0.37 | 2.49 | -2.12 | 28,797 | 22,230 | 0.29 | 15,700 | 17,400 |
| Indonesia (Jakarta) | 1.92 | 1.71 | 0.21 | 1,429,526 | 587,430 | 0.79 | 847,300 | 873,400 |

\* Local GBMSM PSE were generated using GT data in countries' capital.
\*\* Internet access multipliers of 34.1% for Mozambique were applied given limited access in the country.
\*\*\* Extrapolation were conducted based on data from the U.S. Census Bureau in 2025
\*\*\*\* Annual active users of the gay dating app were calculated as 75% of total monthly active user number
^ Due to small survey sample sizes in Senegal (n=23), Ghana (n=61), and Mozambique (n=47), survey multipliers were only applied in Indonesia (n=467). The survey multiplier for Pornhub access was 25.9%; The survey multiplier for Grindr access was 44.8%. Survey adjusted National GTPSE: 5,527,501 (7.41%); Survey adjusted social network app PSE: 1,310,420 (1.76%)

App-based estimates in Ghana (1.34%) and Mozambique (0.29%) exceeded UNAIDS extrapolated figures, while Indonesia and Senegal app-based estimates fell below both GTPSE and UNAIDS values. The largest divergence between methods was observed in Indonesia, where the app-based estimate of 587,430 was less than half the GTPSE of 1,429,526. App-based estimates reflect 2021 monthly active user counts extrapolated to 2025 using national population growth rates. Survey-adjusted sensitivity analysis was conducted for Indonesia only due to small sample sizes in the other three countries (Senegal n=23, Ghana n=61, Mozambique n=47), producing a survey-adjusted GTPSE of 5,527,501 (7.41%) and a survey-adjusted app-based estimate of 1,310,420 (1.76%) (Table 1, Table 2).

**Table 2.** GBMSM estimates from a social network app (Grindr)

|  | Ghana | Indonesia | Mozambique | Senegal |
| --- | --- | --- | --- | --- |
| 2021-12 Monthly Active Users (MAU) | 8507 | 67494 | 909 | 3383 |
| 2021-11 MAU | 9449 | 64071 | 844 | 3193 |
| 2021-10 MAU | 12251 | 65065 | 842 | 3103 |
| 2021-09 MAU | 13425 | 62521 | 808 | 2832 |
| 2021-08 MAU | 13512 | 62286 | 763 | 2835 |
| 2021-07 MAU | 12878 | 59413 | 770 | 2607 |
| 2021-06 MAU | 12332 | 60684 | 737 | 2517 |
| 2021-05 MAU | 12206 | 63162 | 714 | 2539 |
| 2021-04 MAU | 11361 | 61245 | 704 | 2362 |
| 2021-03 MAU | 10784 | 61913 | 696 | 2419 |
| 2021-02 MAU | 10933 | 63501 | 626 | 2308 |
| 2021-01 MAU | 11865 | 68505 | 714 | 2341 |
| Average monthly users | 11625.25 | 63321.67 | 760.58 | 2703.25 |
| Extrapolation for the number of annual users (average*12*0.75) | 104627.25 | 569895 | 6845.25 | 24329.25 |
| Proportion of male with mobile phones that have access to the internet | / | / | 34.10% | / |
| Applying phone procession multiplier | 104627 | 569895 | 20074.05 | 24329 |
| Final Grindr PSE, 2021 | 104627 | 569895 | 20074 | 24329 |
| total population 2021 | 32,372,889 | 275,122,131 | 30,888,034 | 17,462,980 |
| total population 2025 | 35,336,133 | 283,587,097 | 34,206,144 | 19,311,233 |
| population growth rate 2021-2025 | 1.09 | 1.03 | 1.11 | 1.11 |
| Final Grindr PSE, 2025 | **114204** | **587430** | **22230** | **26904** |
| Grindr PSE, %, 2025 | **1.34** | **0.79** | **0.29** | **0.58** |

In sensitivity analyses, estimates based on “gay sex”/”sex” exceeded the extrapolated UNAIDS estimates in all four countries, whereas “anal sex”/”sex” generally produced lower estimates (Supplementary Table 1).

## Discussion

These analyses demonstrate that standardized digital PSE methods can generate cross-country comparable GBMSM estimates in four heterogeneous countries where field-based methods may constrained by criminalization, stigma, and resource availability. The GTPSE generally exceeded UNAIDS estimates, which may reflect underestimation in stigmatizing settings where GBMSM are less likely to be reached through field-based methods or mainstream health services, consistent with prior work on GTPSE(6). According to the UNAIDS 2025 Global AIDS Update, prevention services for key populations had relied heavily on external assistance which was interrupted in early 2025(9). In this context, low-cost internet-derived PSE approaches may be especially valuable for maintaining timely PSE data to support HIV service planning during periods of funding instability.

The social network app PSE were lower in some countries, highlighting variability in platform access settings and geographic differences in concentration of social networks. Social networking apps primarily represent individuals who actively participate in online dating networks, whereas internet search behavior may reflect a broader group of individuals exploring sexual identity or content. These findings indicate that digital platforms could reflect different aspects of GBMSM visibility and online engagement. The divergence between GTPSE and app-based estimates was most pronounced in Indonesia, where the app-based estimate was less than half the GTPSE, likely reflecting uneven platform adoption across a large, geographically dispersed population. In Mozambique, the substantially higher Maputo capital estimate compared to the national figure reflects the concentration of internet access in the capital relative to a national internet penetration rate of 34.1%. These methods are therefore complementary rather than competing, and their divergence is itself informative about the spectrum of GBMSM visibility and online engagement across settings.

The alignment between GTPSE and UNAIDS estimates in Senegal warrants particular attention. Unlike the other three countries in this sample, Senegal had unusually well-developed MSM HIV infrastructure prior to the 2008 criminalization crackdown — including Ministry of Health programs explicitly targeting GBMSM and community-based organizations operating across multiple regions. This institutional engagement likely generated higher-quality underlying data for UNAIDS estimates than is typical in criminalized settings, suggesting the alignment may reflect convergent validity rather than methodological artifact. However, the sensitivity analysis produced a substantially higher estimate in Senegal when “gay sex”/”sex” was used. This raised the possibility that this terminology may better reflect local search behavior than “gay porn”/”porn.” Senegal is also the only Francophone country in the sample, and it is not possible to fully disentangle whether alignment reflects data quality, language effects on search behavior, or both. Future work should test local-language search term equivalents in Francophone settings, consider estimates jointly across multiple search-term combinations and examine whether GTPSE performs differently in countries with more developed MSM data infrastructure.

A key advantage of the approach presented here is methodological standardization. UNAIDS PSE estimates are often derived through varied and potentially incomparable methods across countries challenging cross-country comparisons. The Google Trends and app-based methods applied here use identical procedures across all four settings, producing estimates that are directly comparable regardless of differences in national data infrastructure or political context. This paper extends earlier methodological work lineage demonstrating that Facebook interest targeting and Hornet app data could generate GBMSM PSE consistently higher than UNAIDS estimates across 13 countries. (4) The discontinuation of Facebook’s sensitive audience targeting in 2021-2022 eliminated the broad potential for this approach, and the 2025 interruption of PEPFAR-funded field programs has further constrained resource-intensive in person alternatives. The methods presented here represent a necessary adaptation to this new landscape, demonstrating that reproducible digital PSE remains feasible without dependence on advertising platform data or field infrastructure.

The study has several limitations. Both GT- and app-based approaches rely on assumptions linking online behavior to GBMSM population size and might capture non-GBMSM users. Moreover, GT provides relative rather than absolute search volume and limited transparency regarding its algorithm. Finally, survey multiplier in sensitivity analyses were limited by small survey samples and self-reported measures. App-based estimates reflect 2021 data extrapolated to 2025 as formal data-sharing agreements for public health research with major GBMSM-facing platforms are increasingly difficult to establish access to more current data. A strength of the study is the triangulation of multiple digital data sources to generate GBMSM PSE in settings where conventional approaches may be limited. Although Pornhub audience data were applied to restrict estimates to male viewers, a notable residual limitation is that a proportion of male gay porn viewers identify as heterosexual(10), suggesting GTPSE estimates likely retain an upward bias that may partially account for values exceeding UNAIDS estimates in most settings studied. Moreover, the UNAIDS Key Population Atlas estimates used as comparators are themselves of variable methodological quality and do not represent a validated gold standard, particularly in the four countries studied where criminalization creates structural disincentives for accurate national reporting. Comparisons between GTPSE and UNAIDS figures should therefore be interpreted as triangulation between two imperfect estimates rather than validation of one against the other.”

## Conclusion

These analyses suggested that internet-derived estimates can provide low-cost, feasible, and sustainable GBMSM PSE in low- and middle-income settings. With decreasing external resources, further validating these PSE approaches with additional digital data linkages can support evidence-based allocation of HIV prevention and treatment services even in rights-constrained environments.

## Data Availability

Google Trends data used in this study are publicly available through the Google Trends platform. The aggregated social networking app data are not publicly available because of data-use and confidentiality restrictions but may be available from the corresponding author upon reasonable request.

https://trends.google.com/trends/

## Ethical Considerations

The analyses of Google Trends data and social networking app data were based on publicly available or aggregated, deidentified information and did not involve access to personally identifiable information.

## Consent to Participate

The study was based on deidentified information, therefore consent to participate was not required.

## Consent for Publication

Not applicable. The manuscript does not contain identifiable information relating to individual participants.

## Authors’ Contributions

Y.M., A.G., M.N., S.H., J.H., I.H., and S.B.: Conceptualization and study design. Y.M.: Analysis. Y.M., A.G., M.N., C.H., S.H., J.H., I.H., and S.B.: Drafting article—Revision.

---

**Supplementary Table 1.** Sensitivity analysis comparing national Google Trends–derived GBMSM population size estimates using alternative search terms with UNAIDS estimates.

| Country | “Gay sex”/“sex” GTPSE, n (%) | “Anal sex”/“sex” GTPSE, n (%) | UNAIDS PSE, 2025 |
| --- | --- | --- | --- |
| Senegal | 67,253 (1.44) | 1,955 (0.04) | 58,100 |
| Ghana | 99,471 (1.17) | 78,003 (0.92) | 59,800 |
| Mozambique | 23,038 (0.30) | 47,995 (0.62) | 17,400 |
| Indonesia | 1,335,478 (1.79) | 752,382 (1.01) | 873,400 |

**Abbreviations:** GBMSM, gay, bisexual, and other cisgender men who have sex with men; GT, Google Trends; GTPSE, Google Trends population size estimate; PSE, population size estimate; UNAIDS, Joint United Nations Programme on HIV/AIDS.

